# Types and outcomes of health promotion interventions led by public health students: a scoping review

**DOI:** 10.1101/2023.10.09.23296734

**Authors:** Alejandro Gonzalez-Aquines, Daniel D Bingham, Elizabeth M Kiilu

**Author notes:** Corresponding author: Alejandro Gonzalez-Aquines, The University of Bradford, Richmond Rd, Bradford, England, BD7 1DP.

## Abstract

**Background:** Public health students can contribute to improving population health outcomes; however, this remains an under-researched area. This review aims to assess the extent of the literature related to health promotion interventions led by public health students. This includes what these interventions are and how they are being conducted, as well as their impact on targeted populations.

**Methods:** A scoping review was conducted according to JBI guidelines. A search strategy was developed using the population, context, concept framework. MEDLINE, CINAHL, Embase and Google Scholar were consulted. The selection process involved the screening of titles and abstracts against the inclusion criteria, followed by a full-text screening and data extraction by two reviewers. Disagreements were solved by consensus.

**Results:** 191 studies were identified, but only five of these were included. The target populations included the general public, university students, and minority groups. Students were trained or supervised by experienced staff. The interventions were delivered using diverse means, including innovative approaches through social media. The student-led interventions increased access to preventive services (screening, vaccinations) and improved health knowledge within the target groups.

**Conclusion:** Public health students make impactful contributions by increasing both the access to and the use of preventive services, as well as by promoting health knowledge. Our study contributes to the professionalisation of public health, highlighting for the first time the role of public health students in improving health outcomes. Furthermore, our study sheds light on the students’ impact on the reduction of health inequalities, particularly amongst minority groups.

## Introduction

Student-led interventions can be a sustainable and cost-effective approach to delivering targeted public health messages. These messages can be delivered through community health clinics, mass campaigns, or other types of health education (1). Another advantage of student-led interventions is the ability to increase access to health promotion and disease prevention services at community level (2). Hence, student-led interventions contribute to ensuring that underserved populations (such as uninsured patients or minority groups) can still access health promotion interventions, whilst giving the students themselves valuable opportunities to practice what they learn in real-world settings.

Student-led health interventions have been shown to produce desirable health outcomes (1). However, the available literature is generated mainly by interventions led by students in areas such as pharmacy (3), nursing (4), dentistry (5), and medicine (6, 7), with limited information from public health students. The limited literature on public health student-led interventions might be linked with the relatively recent professionalisation of public health. In the United Kingdom, for example, the Public Health Registry was founded in 2003, while the General Medical Council dates all the way back to 1858 (8, 9). Similarly, the Association for Schools of Public Health in the European Region, in collaboration with the World Health Organisation, only published the roadmap to professionalising the public health workforce in Europe as recently as 2022 (10).

Both experienced public health professionals and those still in training can significantly contribute to improving a population’s health. In the aftermath of the COVID-19 pandemic, this has become a major focus for governments and health authorities the world over (11, 12). Public health involves the protection of populations from disease/injury, the prevention of illness, and the promotion of health (13). The monicker of ‘core public health professionals’ refers to those graduating with a degree (bachelors or postgraduate) in public health (10). Such professionals are trained in a diverse range of competencies. These include health protection/promotion, the prevention, monitoring and evaluation of disease, and even leadership abilities (10). The knowledge and skills acquired during undergraduate and postgraduate programmes in public health equip future professionals with the necessary tools to make important contributions to a population’s health. However, there is limited information about how these skills are applied during public health professionals’ training years, as well as the impact of student–led public health interventions.

As evidence shows, students from other healthcare professions are already applying their knowledge and assisting local populations during their training years. Thus, we wondered what the available literature was regarding public health student-led health promotion campaigns. A preliminary search of MEDLINE, the Cochrane Database of Systematic Reviews, and JBI Evidence Synthesis was conducted. From this search, no current systematic reviews or scoping reviews on the topic were identified as being underway. Therefore, this scoping review aimed to assess the extent of the literature related to public health student-led health promotion interventions. We set out to examine the nature of these interventions, the methodology behind them, and their ultimate impact on the target populations.

## Methods

The protocol of the scoping review was registered on the Open Science Framework (14). The rest of the methods are presented according to the PRISMA-ScR (15).

### Eligibility criteria

The eligibility criteria was defined based on the participants, concepts, and context (PCC) framework. Participants were defined as public health students registered in undergraduate or postgraduate programmes. Students from other healthcare programmes (i.e., nursing, medicine, paramedics) were excluded. The concept referred to student-led health promotion interventions. These interventions could have occurred with or without supervision. Similarly, interventions both with and without prior training were included. Interventions where students merely shadowed a trained healthcare professional were not included. The context was defined as interventions developed under clinical and non-clinical settings, regardless of geographical location. Finally, the types of sources included qualitative, quantitative, and secondary research designs. Text and opinion papers were not considered for inclusion in this scoping review, unless they presented original quantitative or qualitative data addressing the inclusion criteria of this review. Only studies published in English were included.

### Information sources

An initial limited search of MEDLINE and CINAHL was undertaken to identify relevant articles. The text words in the titles and abstracts of these articles, as well as the index terms describing the content, were used to develop a full search strategy (see **Supplementary Material**). The search strategy, including all identified keywords and index terms, was adapted for each database and/or information source. The reference list of all included sources of evidence was screened for additional studies. No limitations were added to the search strategy. The databases to be searched included MEDLINE, CINAHL and Embase. Literature, including grey literature, was searched for in Google Scholar using the same key terms as the rest of the databases. Results from Google Scholar were limited to the first ten pages, and a result list was generated using Publish or Perish (16).

### Selection of sources of evidence

Following the search of existing studies, all identified references were collated and uploaded into Covidence (17), with any duplicates being removed. The selection process featured the screening of titles and abstracts against the inclusion criteria, followed by full-text screening and data extraction.

Each of these steps was performed by at least two reviewers. Disagreements were resolved via consensus or by a third reviewer. Reasons for excluding sources of evidence in full texts that do not meet the inclusion criteria are reported using the PRISMA flow diagram below (15).

### Data charting process

Data was extracted by at least two independent reviewers using a data extraction tool developed by the reviewers. The extracted data included specific details about the settings and types of students leading the interventions, as well as the interventions’ scope, training, implementation, and outcomes. A pilot of the extraction form was conducted using the first two studies, with no major modifications. An additional option was only added when specifying if the programme was an undergraduate, postgraduate degree, or both.

### Synthesis of the results

The analysis was limited to a description of the interventions, which reported the elements mentioned in the data extraction form. A narrative summary of the findings will accompany the tabulated results, which will describe how these results relate to the objective and question of the review.

## Results

A total of 191 studies were identified from the searched databases. After duplicates were removed, 173 titles and abstracts were screened and only five studies were included following the full text screening process (**Figure 1**). All of the studies were conducted in the United States of America and most of them described interventions led by undergraduate students (**table 1**).

**Figure 1.**
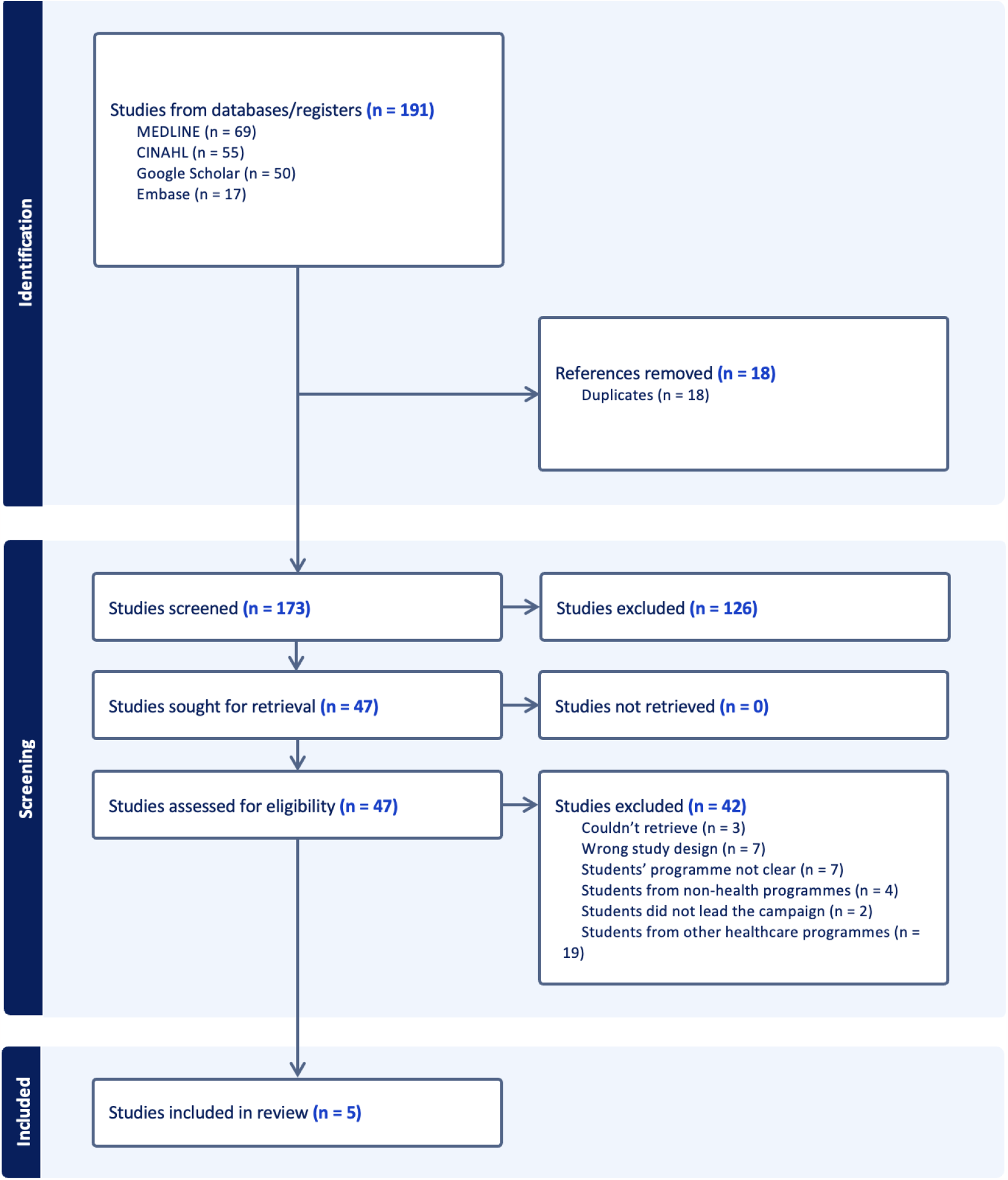
Flow diagram of the included studies. Elaborated from Covidence (17).

**Table 1.**
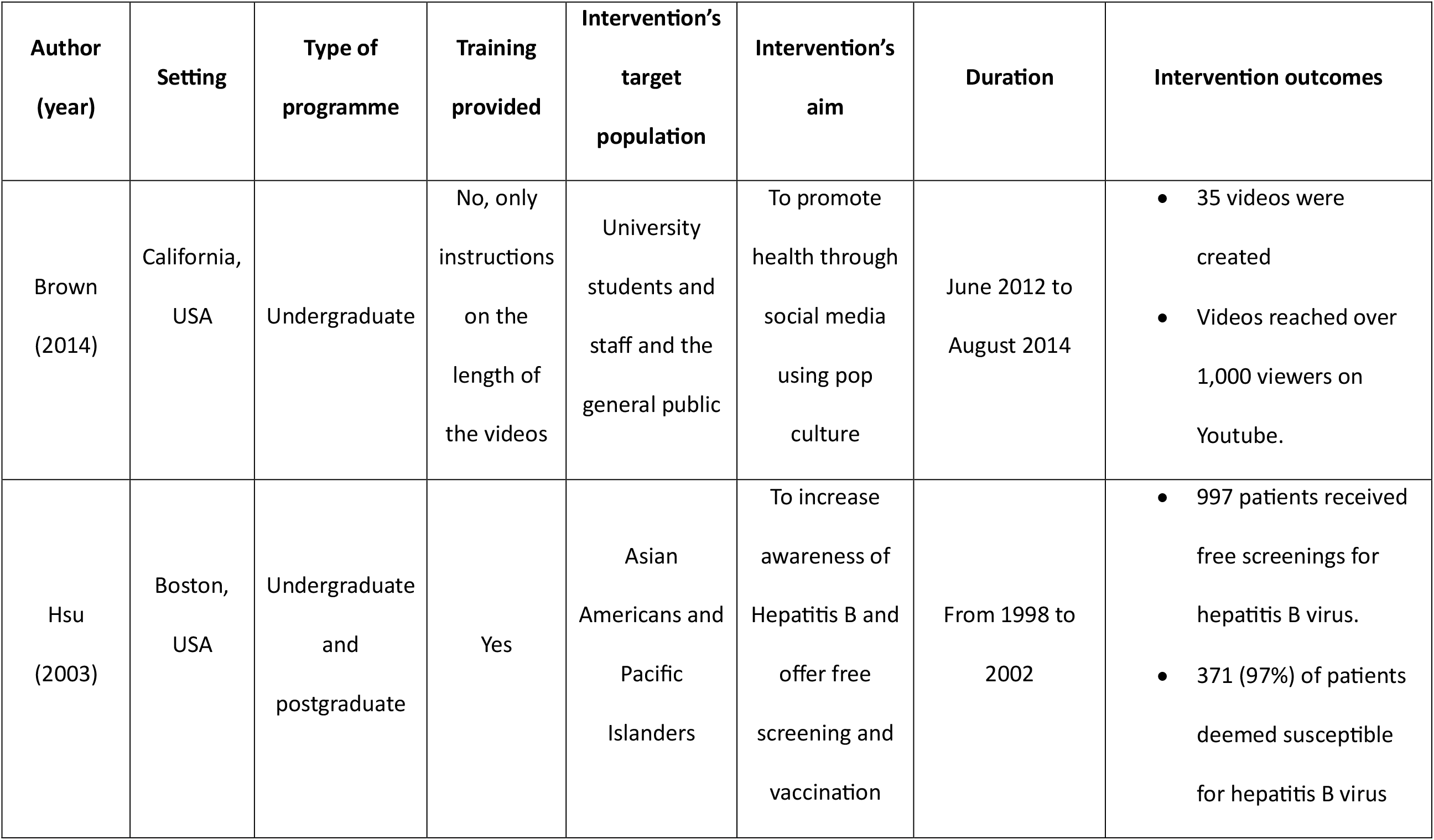
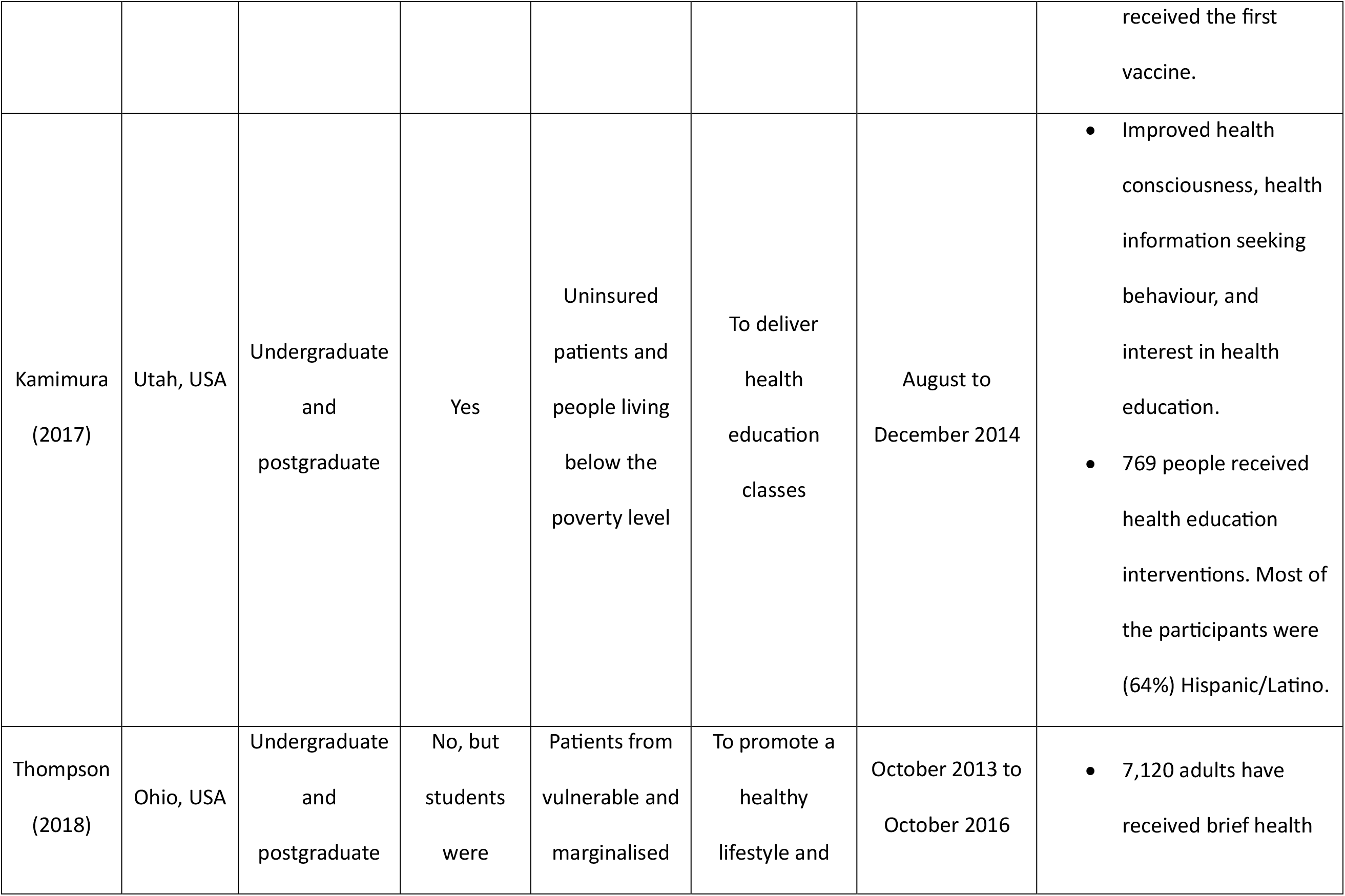
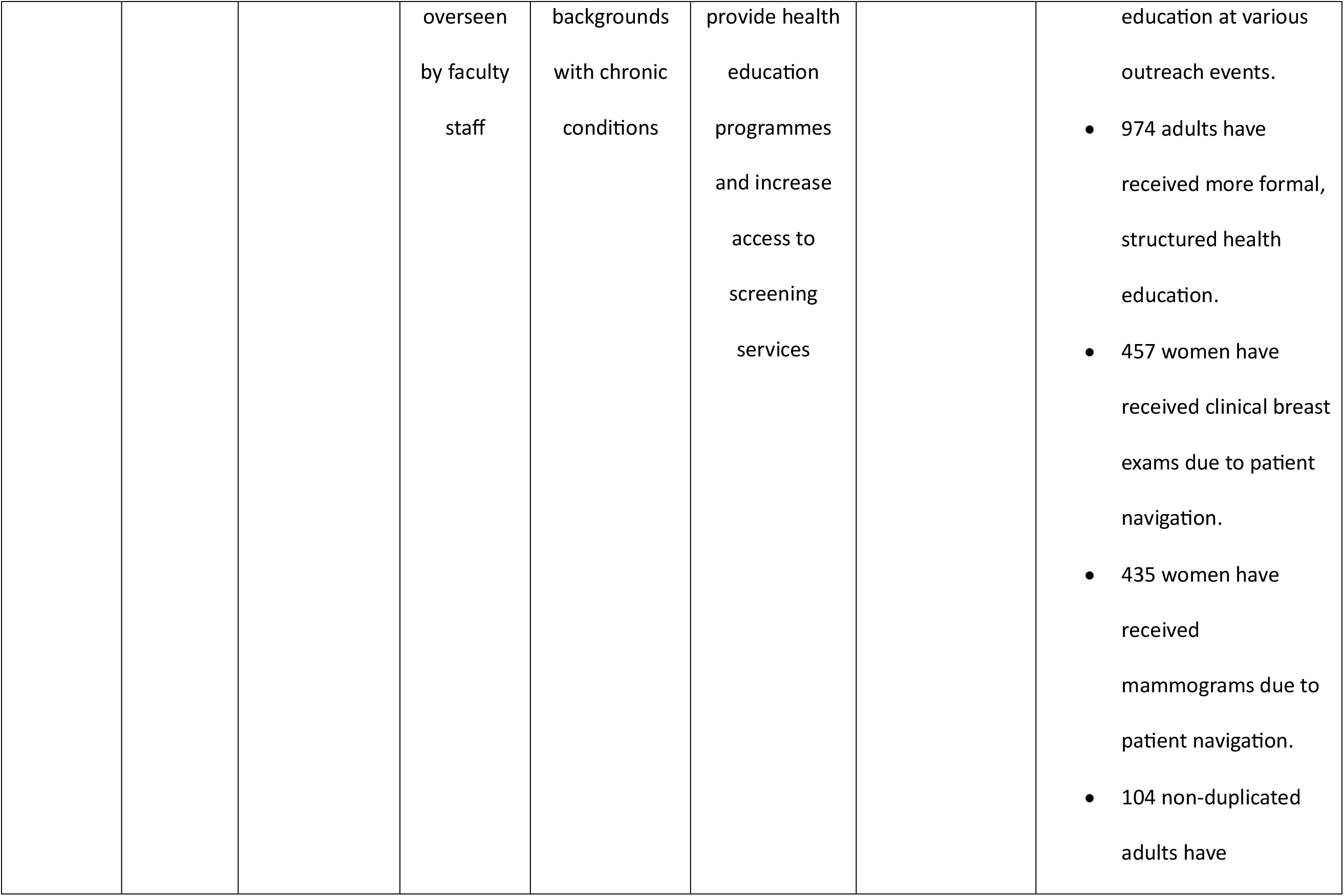
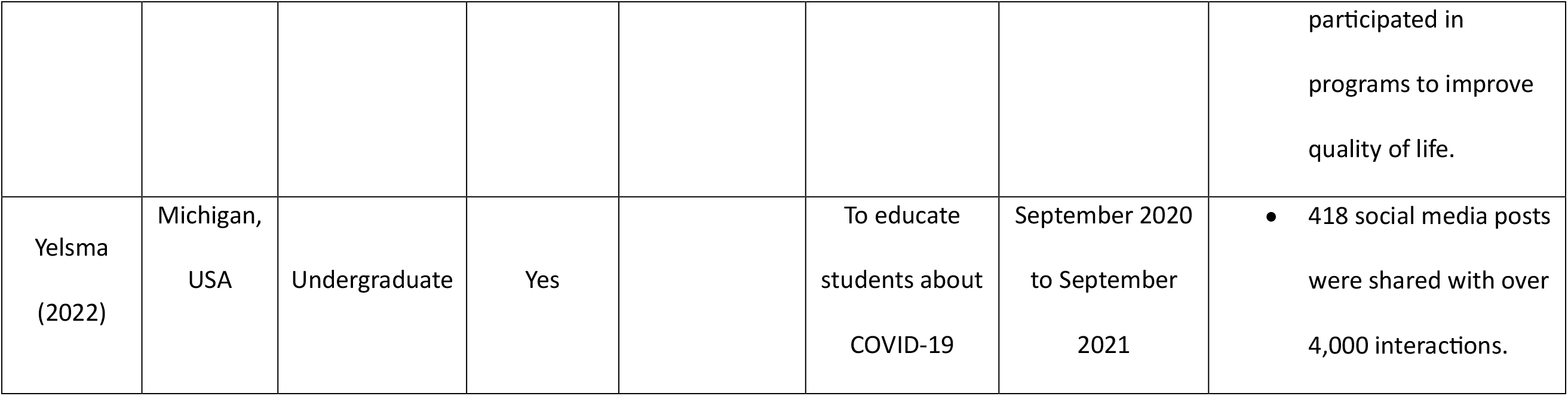
Summary of the included studies.

| Author<br>(year) | Setting | Type of<br>programme | Training<br>provided | Intervention's<br>target<br>population | Intervention's<br>aim | Duration | Intervention outcomes |
| --- | --- | --- | --- | --- | --- | --- | --- |
| Brown<br>(2014) | California,<br>USA | Undergraduate | No, only<br>instructions<br>on the<br>length of<br>the videos | University<br>students and<br>staff and the<br>general public | To promote<br>health through<br>social media<br>using pop<br>culture | June 2012 to<br>August 2014 | 35 videos were<br>created<br>Videos reached over<br>1,000 viewers on<br>Youtube. |
| Hsu<br>(2003) | Boston,<br>USA | Undergraduate<br>and<br>postgraduate | Yes | Asian<br>Americans and<br>Pacific<br>Islanders | To increase<br>awareness of<br>Hepatitis B and<br>offer free<br>screening and<br>vaccination | From 1998 to<br>2002 | 997 patients received<br>free screenings for<br>hepatitis B virus.<br>371 (97%) of patients<br>deemed susceptible<br>for hepatitis B virus<br>received the first vaccine. |
| Kamimura (2017) | Utah, USA | Undergraduate and postgraduate | Yes | Uninsured patients and people living below the poverty level | To deliver health education classes | August to December 2014 | Improved health consciousness, health information seeking behaviour, and interest in health education.<br>769 people received health education interventions. Most of the participants were (64%) Hispanic/Latino. |
| Thompson (2018) | Ohio, USA | Undergraduate and postgraduate | No, but students were | Patients from vulnerable and marginalised | To promote a healthy lifestyle and | October 2013 to October 2016 | 7,120 adults have received brief health |
|  |  |  | overseen<br>by faculty<br>staff | backgrounds<br>with chronic<br>conditions | provide health<br>education<br>programmes<br>and increase<br>access to<br>screening<br>services |  | education at various<br>outreach events.<br>• 974 adults have received more formal, structured health education.<br>• 457 women have received clinical breast exams due to patient navigation.<br>• 435 women have received mammograms due to patient navigation.<br>• 104 non-duplicated adults have<br>participated in<br>programs to improve<br>quality of life. |
| Yelsma<br>(2022) | Michigan,<br>USA | Undergraduate | Yes |  | To educate<br>students about<br>COVID-19 | September 2020<br>to September<br>2021 | 418 social media posts were shared with over 4,000 interactions. |

The target populations of the health promotion interventions varied widely. They included the general public (18), university students (18, 19), and minority groups (20-22). All interventions that occurred in clinical settings targeted minority groups, such as Asian American and Pacific Islander (20), patients attending a free clinic (21), and people from vulnerable and marginalised backgrounds (i.e., transgender women and uninsured adults) (22). Two of the interventions were carried out through social media, using such methods as interacting with Instagram users (19) and creating videos to share information and inform the general public about different conditions (18).

Training was provided for most of the interventions. When training was not provided, a set of instructions was given or a senior member of staff was allocated to the project, depending on the type of intervention. For example, in the creation of videos described by Brown et al, students were simply provided with a set of instructions on how to develop their own video content. On the other hand, when training students to deliver health education to patients attending a free clinic, a coach was assigned to guide the students on the delivery of the intervention (22).

Impact was measured in different ways, depending on the intervention delivery mode. Social media interventions used metrics such as the number of viewers (18) or content created and interactions (i.e., likes) (19). Interventions carried out in clinical settings measured the number of participants receiving screenings (20, 22), health education (21, 22) and vaccinations (20).

## Discussion

Our study identified literature describing the implementation of health promotion interventions led by public health students. The types of intervention varied widely, from social media-based initiatives to more traditional efforts occurring in clinical settings. This demonstrated the diverse set of skills acquired by public health students. The interventions mainly involved disadvantaged populations, and led to an increase in screenings and the use of services to prevent and promptly identify serious conditions (such as breast cancer and hepatitis B).

Public health students led health promotion interventions using diverse means. Traditional face-to-face contact was prevalent across the studies, but the use of social media platforms to promote health was also reported in certain cases (18, 19). Due to the nature of the profession, communication is one of the core elements that public health students need to develop. This includes the sharing of health information with people from different backgrounds, including policy makers and people from minority groups.

Our findings align with the Agency for Public Health and Education Accreditation (APHEA) requirements for accreditation, which include competencies in communication, advocacy and marketing (23). The Association of Schools and Programmes of Public Health (ASPPH) also includes advocacy and communication through mass media as part of the core curriculum for public health programmes (24). In contrast to medicine (25) and nursing (26), the focus of communication training for public health students is geared towards the wider population, rather than individual patients and family members. Moreover, by engaging in advocacy training, public health professionals gain the skills to ensure that voices from disadvantaged groups are heard by policy makers and relevant stakeholders. This is something that was captured in this review.

The included studies conducted in clinical settings targeted minority and disadvantaged groups. It is not surprising that the focus of these interventions was on underserved populations, as the foundational values of public health revolve around equality, social justice and protecting the health of communities and individuals. In other words, working towards identifying and tackling health inequalities in societies (27). Lee et al (28) discuss the values of public health and conclude that health is essential for human flourishing, and that liberal governments must provide equal opportunities for all members of society to access healthcare (28). These values are also reflected in official frameworks for public health professionals, for instance the Public Health Skills and Knowledge Framework (PHSKF) in the United Kingdom (29). The PHSKF is divided in three main areas: technical, context, and delivery. Each of these areas is subdivided into functions where the main focus is on identifying and addressing health inequalities, as well as assuming leadership and ensuring efficient resource management.

In addition to focusing on health promotion, the included studies showed that student-led interventions improved screening and the early detection of chronic conditions. The role of students in screening services is not new; there is a vast amount of literature showing that medical students can use screening services to increase the early detection of diseases, such as hypertension, diabetes, and cancer (2, 30-32). However, the unique focus of our findings is that public health students in particular – upon receiving adequate training or supervision – can also engage in these types of activities. This is particularly relevant, considering that many health services are starting to shift from focusing on hospital and acute care to enhancing preventive care (33). Public health professionals can therefore take a more active role in the screening of noncommunicable diseases that represent significant causes of death worldwide.

It is also important to mention that most of the activities were conducted under multidisciplinary settings, emphasising the role of collaboration between clinical and non-clinical professionals. Lastly, although not a primary focus of this review, one of the full-text reviewed articles focused on satisfaction from student-led campaigns. This article stated that more than 80 percent of respondents expressed that the student-led activities had a positive impact on their health and well-being, while over 90 percent advocated the importance of students leading health promotion activities (34). These findings all indicate that public health students can indeed lead health promotion campaigns to increase awareness, education and screening. Furthermore, these campaigns are positive experiences for the recipients and culminate in desirable outcomes.

Our study was not exempt from limitations. While the research team used the most relevant databases to identify studies related to the review’s aim, we did also limit our search to studies published in English. Not surprisingly, most of the included studies were from the United States of America, where public health programs are most prevalent when compared to other English-speaking countries (35). Incorporating studies published in other languages could have increased the sample size, particularly as public health schools from non-English speaking countries have a long legacy of public health education. For example, the School of Public Health of Mexico has been active for well over a century (36).

Another limitation was the focus on health promotion. While this is one of the main duties of public health professionals, it is not the only role they play. By focusing on health promotion, we excluded tasks such as advocacy campaigns and health policy, which could have provided further insight into how public health students are engaged in such activities.

This study investigated the role and impact of public health students in leading health promotion interventions. This is the first time a review has explored this particular topic, adding an important contribution to both the literature and the professionalisation of public health. We described how public health students engage with underserved communities, increase access to preventive care, and use diverse means to communicate essential information in the quest to promote healthy lifestyles.

Further primary research is needed to have a better understanding of the impact of public health students’ actions, with the outcomes measured using standardised tools. Nonetheless, our study represents another step forward on the path towards professionalising public health education. It demonstrates that engaging students in health promotion can benefit the most vulnerable, leading to tangible reductions in health inequalities.

## Funding

No funding was received for the development of this research.

## Data Availability

All data produced in the present work are contained in the manuscript

## Acknowledgements

The authors thank Alex Palmer for his editorial services. The authors also thank the Faculty of Health Studies at the University of Bradford for providing access to Covidence systematic review software, Veritas Health Innovation, Melbourne, Australia..

## Conflicts of Interest

None declared.

### Key points

- Student-led interventions are an efficient way to reach the local population.
- Public health students are trained to enhance population health outcomes.
- Interventions led by public health students are shown to improve access to preventive services, particularly among minority and underserved groups.
- Increasing the prevalence of public health student-led interventions can contribute to reducing health inequalities.

## Notes

### Competing Interest Statement

The authors have declared no competing interest.

### Funding Statement

This study did not receive any funding

