## Supplementary Material for "Types and outcomes of health promotion interventions led by public health students: a scoping review"

**S1: Search strategy development**

| PCC | Search terms |
|---|---|
| Population | Public health students |
| Concept | Student-led OR student-delivered OR student-run |
| Context | Health education OR health promotion OR campaign* OR health intervention |

*The search strategy will combine the terms for each PCC element using AND.*

Example of MEDLINE via EBSCO search strategy

| PCC | Search terms |
|---|---|
| S1 | MH "Public Health+" |
| S2 | Student-led |
| S3 | student-delivered |
| S4 | student-run |
| S5 | MH "Health Education+" |
| S6 | MH "Health Promotion+" |
| S7 | health campaign |
| S8 | Combines S2, S3, S4 using OR |
| S9 | Combines S5, S6, S7 using OR |
| S10 | Combines S9 with S10 using AND |
| S11 | Combines S11 with S1 using AND |
